# Selective IgA_2_ deficiency in a patient with small intestinal Crohn’s disease

**DOI:** 10.1101/2022.12.03.22282315

**Authors:** Pablo Canales-Herrerias, Yolanda Garcia-Carmona, Hadar Meringer, Gustavo Martinez-Delgado, Michael Tankelevich, Jean-Frederic Colombel, Charlotte Cunningham-Rundles, Andrea Cerutti, Saurabh Mehandru

## Abstract

The human IgA response is composed of two structurally different subclasses termed IgA_1_ and IgA_2_. Compared to IgA_1_, IgA_2_ has a shorter hinge region, which makes it more resistant to bacterial proteases. IgA_1_ is produced both systemically and in mucosal surfaces, whereas IgA_2_ is mostly confined to the intestines. While the overall IgA response is known to be involved in intestinal homeostasis, the specific contribution of IgA_1_ and IgA_2_ remains largely unknown (Chen 2020). Selective IgA deficiency (SIgAD) is the most prevalent primary immune deficiency. About half of SIgAD cases are associated with heterogeneous but generally mild clinical manifestations. Anecdotal evidence of IgA_2_ deficiency is available (van Loghem 1983, Ozawa 1986, Engström 1990), however no associations with clinical manifestations have been reported.

Here, we describe the occurrence of a selective IgA_2_ deficiency in a patient (CD068) with small intestinal Crohn’s disease (CD). The patient had undetectable IgA_2_^+^ cells and secreted IgA_2_ antibody in both intestine and circulation. Among other features, patient CD068 presented with duodenal and ileal inflammation. To our knowledge, this is the first case of IgA_2_ deficiency with a potential link to IBD, which might shed new insights into potential IgA_2_-specific functions.

## Results

Small intestinal (ileal) and colonic biopsies were obtained from patient CD068 (male, 36-40 y/o) with history of duodenal, ileal, and perianal Crohn’s Disease (CD). A cohort of healthy donors (n=22) and patients with IBD (n=40) served as controls (Tables S1-S2). Mucosal mononuclear cells and peripheral blood mononuclear cells (PBMCs) were characterized by flow cytometry (Figure 1A).

**Figure 1.**
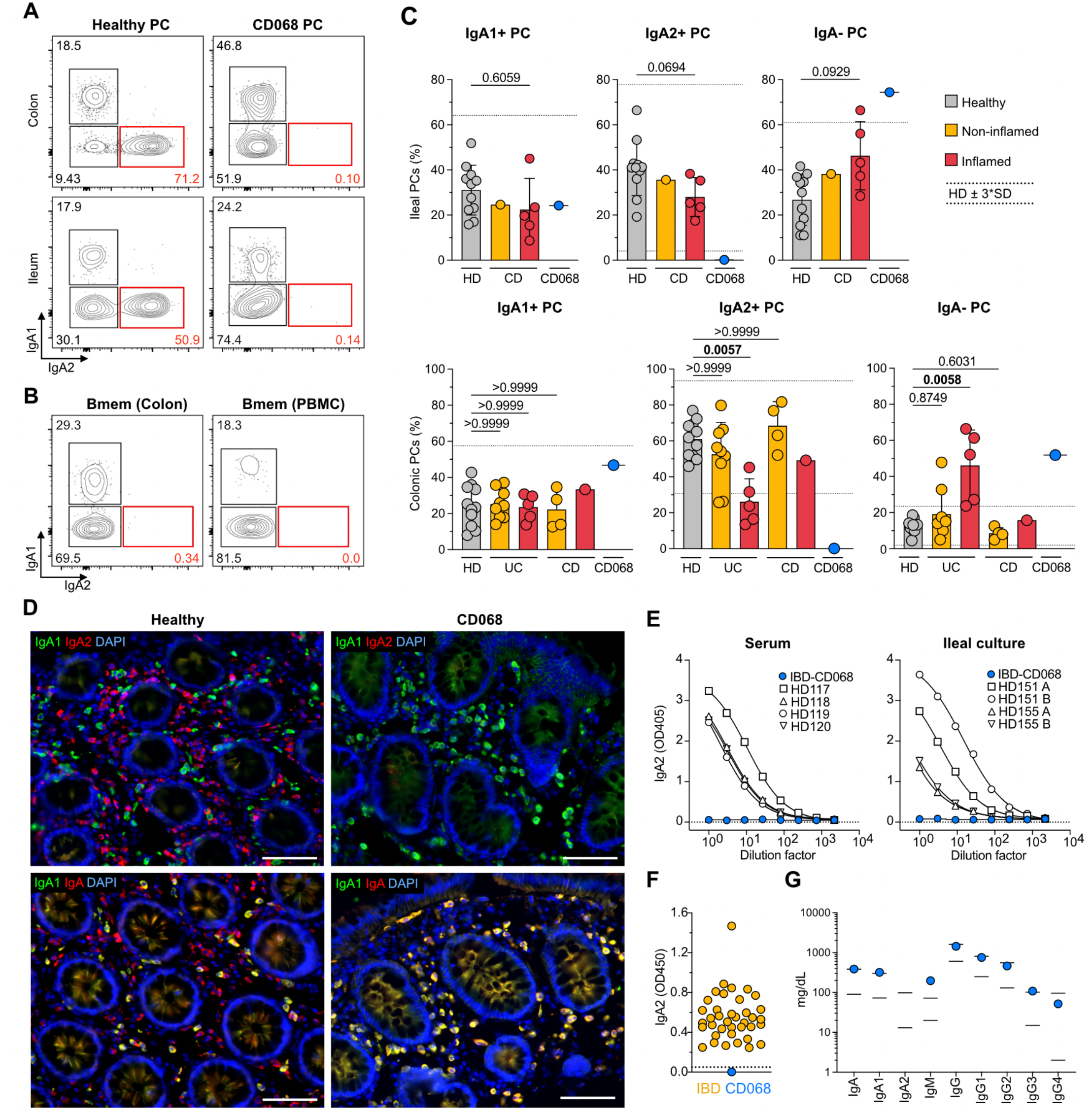
Selective IgA_2_ deficiency in a patient with small intestinal CD. (A) Flow cytometric staining of IgA_1_ and IgA_2_ on intestinal lamina propria and circulating PCs from a representative control HD (left) and IBD patient CD068 (right). (B) Flow cytometric staining of IgA_1_ and IgA_2_ on memory B cells from colonic lamina propria (left) and peripheral blood (right) from IBD patient CD068. (C) Frequency of PCs from the mucosa of control HDs (gray), noninflamed IBD patients (yellow), inflamded IBD patients (red), and IBD patient CD068 (blue symbol). Ileal mucosal samples are represented in top panels and Colonic mucosal samples in lower panels. Mean and SD are shown. Interval estimate calculated as 3 X SD around the mean of control HDs. (D) Immunofluorescence staining of IgA_1_ and IgA_2_ (top) as well as IgA_1_ and total IgA (bottom) from a representative control HD (left) and IBD patient CD068 (right). (E) ELISA of secreted IgA_2_ from serum and ileal tissue culture supernatant of control HDs and IBD patient CD068 (blue symbol). (F) ELISA of secreted IgA_2_ from plasma of UC and CD patients (yellow), and IBD patient CD068 (blue symbol). (G) Nephelometry of total serum immunoglobulin levels from control HDs (black lines indicate normal range) and IBD patient CD068 (blue symbol).

A complete absence of IgA_2_^+^ plasma cells (PCs) was noted in the colon, ileum and peripheral blood of CD068, while IgA_1_^+^ PCs were present throughout (Figures 1A, S2). Further, among CD19^+^IgD^−^IgM^−^CD27^+^ memory B cells, IgA_2_-switched cells were absent in the ileum, colon and PBMCs of CD068 (Figure 1B). To compare PCs from CD068 with those from additional IBD patients, we obtained colonic and ileal biopsies from a cohort of 8 UC patients, 6 CD patients, and 19 healthy donors (HDs), and performed flow cytometry on isolated cells (Figure 1C). When intestinal PC populations were analyzed, the frequency of ileal and colonic IgA_1_^+^ PCs was comparable in IBD patients and HDs. Similarly, the frequency of IgA_1_^+^ PCs from CD068 fell within the interval estimate for HDs (3 standard deviations around the mean). Conversely, a significant loss of IgA_2_^+^ cells was detected in inflamed colonic areas from UC patients compared to HDs, which suggests that IgA_2_ could have subclass-specific homeostatic functions. Although not significant, there was a trend for the loss of IgA_2_^+^ cells in inflamed ileal tissues from CD patients. Together with the loss of IgA_2_^+^ PCs, we found an expansion of non-IgA PCs in the same inflamed areas of ileum and colon of IBD patients compared to HDs, as suggested by previous reports (Castro-Dopico 2019, Uzzan 2022). The frequency of IgA^−^ PCs in CD068 was above the HD interval estimate in the ileum, consistent with the presence of inflammation in this tissue. Interestingly, a strong increase of IgA^−^ PCs was also seen in the uninflamed colon of CD068.

To confirm the loss of IgA_2_^+^ cells in the gut mucosa, we used tissue immunofluorescence (IF) to stain for IgA_1_^+^ and IgA_2_^+^ cells. We did not find any IgA_2_^+^ cells in the intestinal mucosa of CD068, while IgA_1_^+^ cells were readily detected (Figure 1D). To exclude the possibility of lack of detection by our primary antibody, we stained tissue for IgA_1_ and total IgA, and found that all IgA^+^ cells were also IgA_1_^+^ in CD068. Next, we asked whether secreted antibodies lacked IgA_2_ in patient CD068 and measured the concentration of serum IgA_2_ by ELISA. While IgA_2_ could be readily measured in HDs, no detectable IgA_2_ was found in patient CD068 (Figure 1E). In addition, we compared circulating IgA_2_ from CD068 with a cohort of 39 IBD patients (27 CD and 12 UC), and found detectable IgA_2_ in the entire cohort, except CD068. Considering that IgA_2_ is mostly produced in the intestinal mucosa, we also measured gut IgA_2_ secretion. For this purpose, we used a biopsy culture system to harvest tissue-derived antibodies (Mesin 2011). We found no detectable IgA_2_ in biopsy culture supernatant from patient CD068, while we could detect it in biopsy culture supernatant from 4 HD samples. Furthermore, we measured the IgA coating profile of fecal microbiota, and found no IgA_2_-coated bacteria in CD068, while IgA_2_ coating was readily detected in HD microbiota (Figure S1). To establish if this phenotype was specific to IgA_2_, we measured each antibody class and subclass in the serum from patient CD068. In contrast to IgA_2_, which remained undetectable, the other antibody isotypes were detectable and comparable to the levels seen in HDs. Interestingly, circulating IgM from patient CD068 was more abundant than IgM from HDs, suggesting that the loss of IgA_2_ was compensated for by IgM.

Next, we examined whether B cells from patient CD068 did not produce IgA_2_ due to an intrinsic B cell defect and stimulated the patient’s PBMCs *in vitro* for 10 days using CD40L and IL-2. While HDs induced both IgA_1_^+^ and IgA_2_^+^ PCs, no IgA_2_^+^ PCs were induced by patient CD068 (Figure S2). Furthermore, a 407 gene panel for immune deficiency was used to examine genetic alterations (data not shown). No homozygous alterations were noted. The patient was found to have 7 variants of unknown significance in *AP3B1, C7, CCBE1, ITGB2, SAMD9 TCF3*, and *USB1*, and one pathogenic recessive mutation in *TCIRG1* (associated with autosomal recessive osteopetrosis).

## Discussion

IgA is thought to be central to the maintenance of intestinal homeostasis and prevention of inflammation. Selective IgA deficiency (SIgAD) is often asymptomatic. Nevertheless, the prevalence of inflammatory bowel disease, especially CD, is higher among the SIgAD population (Ludvigsson 2014). Although patient CD068 did not lack IgA_1_, there may be common mechanisms in the pathogenesis of SIgAD and IgA_2_-deficiency-related inflammation, considering that IgA_2_ is particularly abundant in the intestine.

Selective loss of IgA_1_ associated with loss of IgG_2_, IgG_4_ and/or IgE was described in a few patients due to large homozygous gene deletions in the *IgCH* gene (van Loghem 1983). Loss of IgA_2_ in serum was first reported in two members of one family (Zegers 1986); this isotype was also not expressed on lymphocytes even after B cell activation *in vitro*. Using genetic marker (Gm) allotyping, deletion of the *□2* gene was proposed (Zegers 1986), however no gross deletions or rearrangements could be detected in these subjects (Oliviero 1986). Loss of IgA_2_ by gene deletion was later documented in one patient, who also had loss of IgG_2_ and IgG_4_ due to homozygous deletions of the *□2* and *□4* genes (Bottaro 1989, Engstrom 1990). Patient CD068 had normal expression of IgG subclasses confirming no loss of these genes and suggesting a more restricted genetic loss or another mechanism for lack of IgA_2_.

Although the functional differences between IgA_1_ and IgA_2_ remain unclear, these IgA subclasses possibly serve unique roles in the gut mucosa. For example, given its higher resistance to bacterial proteolysis, IgA_2_ may be particularly important for preventing luminal microbes from reaching the epithelium, in a process called immune exclusion. Should this be the case, the lack of IgA_2_ could increase the penetration capacity of commensal microbes across the gut epithelium. In support of this idea, a loss of IgA_2_^+^ PCs in inflamed IBD tissue has been described previously (Kett 1987 & 1988, Boland 2020). The resulting pro-inflammatory environment would favor the induction of IgG^+^ PCs. Accordingly, an expansion of IgG^+^ PCs has been recently described in inflamed IBD tissue (Uzzan, 2022). Of note, IgG released by these pathological PCs would contribute to gut inflammation by delivering FcR-mediated pro-inflammatory signals to myeloid cells (Castro-Dopico 2019).

The small intestine is the largest reservoir of IgA^+^ PCs in the body (Bunker 2015) and contains a bacterial community that is more heavily coated by IgA than in the large intestine (Sterlin 2019). These properties could render the small intestine more susceptible to tissue injury in the context of impaired IgA production. Consistent with this possibility, mouse IgA deficiency has been linked to spontaneous inflammation of the small intestine (Nagaishi 2022); this inflammation was associated with dysbiosis of the local microbiota, which encompassed expansion of segmented filamentous bacteria. Similarly, human IgA_2_ deficiency might unbalance the composition and/or function of the gut microbiota, leaving the small intestine particularly susceptible to inflammation. Although a causal relationship between the lack of IgA_2_ and CD remains unproven, our study shows novel evidence that documents an association between IgA_2_ deficiency (confirmed by multiple, orthogonal assays) and small bowel CD with atypical features (duodenal inflammation). Further studies aimed at dissecting the specific function and reactivity of gut IgA_1_ and IgA_2_ could lead to a better understanding of the contribution of IgA subclasses to IBD pathogenesis.

## Data Availability

All data produced in the present study are available upon reasonable request to the authors

## Author contributions

PC, SH and AC designed research. PC, YG, HM and GM performed research. PC analyzed data. PC, SM and AC wrote the manuscript.

## Acknowledgments

We thank the patients who participated in the study. This work was supported by the following grants: Anonymous Donor (SM and JFC), NIH/NIDDK R01 123749 (SM).

## Materials and Methods

### Patients

Patients were recruited between 2019 and 2022 in the Inflammatory Bowel Disease Center, the Gastroenterology Department and the Digestive Endoscopy Unit at Mount Sinai Hospital, NY. In accordance with the institutional review board, the protocol was approved and informed consent was obtained from all patients. Detailed patient characteristics are included in Table S1 and S2. Patients enrolled in the study were asked to donate intestinal biopsies and blood. Biopsies were collected during colonoscopies planned for regular care.

Patient CD068 (male, 36-40 y/o) had abdominal pain and bleeding at age 16-20 and diagnosed with stricturing Crohn’s disease. He had no medical history of respiratory tract infections, illnesses or hospitalizations. One parent has a history of pemphigus vulgaris; there was no history of consanguinity.

All patient IDs are internal to our laboratory and are used only for purposes of this research project.

### Intestinal tissue processing

During colonoscopy, 8-12 biopsies from non-inflamed and/or inflamed regions of the descending colon and terminal ileum were collected with forceps directly into ice-cold RPMI and transported to the laboratory. All biopsies were processed within 3h of collection. Two consecutive rounds of dissociation were performed to remove the epithelium, each consisting of 20 min incubation (37°C, 215rpm) in 10ml HBSS (no calcium and magnesium) (Gibco) containing 5mM EDTA (Invitrogen), 100mM Hepes (Lonza) and 10% FBS, followed by 30s vortex and washing with HBSS. Biopsies were then transferred into 10ml of digestion media, made of RPMI (Gibco) containing 0.5mg/ml Collagenase IV (Sigma-Aldrich), 0.1mg/ml DNAse-1 (Roche) and 1% FBS, and were incubated for 40 min (37°C, 215rpm). Biopsies were mechanically disrupted using a syringe needle, before two filtering and washing steps (RPMI) through 100μm and 40μm cell strainers. The single cell suspension was centrifuged (400G, 10 min) and resuspended in RPMI for downstream analysis.

### Blood processing

Blood was drawn in EDTA tubes, transported to the lab at room temperature, and processed within 3h of collection. Blood was diluted with PBS 1X (Gibco) and gently overlaid on lymphocyte separation medium (MP Biomedicals). After centrifugation, mononuclear cells were recovered and washed two times with PBS. Serum was collected and immediately stored at −80°C.

### Flow cytometry

After isolation from tissue, cells were incubated for 20min at 4°C in PBS containing a mix of staining antibodies. Cells were then washed with PBS and fixed using PBS with 2% formaldehyde (ChemCruz) for 20 min, before washing with PBS. Cells were resuspended in PBS and acquired within 24h using LSR Fortessa (BD Bioscience). Data was analyzed using FlowJo v10 (FlowJo). A list of antibodies is displayed in Table S3.

### Tissue immunofluorescence

Immunofluorescence staining was performed on formalin-fixed, paraffin-embedded intestinal tissue sections of 5μm thickness. Samples were deparaffinized through two 5 min xylene baths, followed by consecutive washes in solutions of decreasing concentration of ethanol in distilled water (100, 90, 80 and 70%), and two 5 min baths in PBS. Antigen retrieval was done using Dako antigen retrieval solution (Dako) and 15 min heating in pressure cooker, before washing 3x with PBS. Samples were blocked using 10% goat serum (Life technologies) or donkey serum (Southern Biotech) for 1h at room temperature. Primary antibodies were then added to the tissue in serum, and samples were incubated overnight at 4°C. Slides were washed with PBS, before adding secondary antibodies and DAPI, which were incubated for 1h at room temperature. Slides were washed with PBS and mounted using Fluoromount G (Electron Microscopy Sciences). A list of antibodies is included in Table S3. Tissue was visualized using a Nikon Eclipse Ni microscope and a Nikon DS-Qi2 camera.

### ELISA

Maxisorp ELISA plates (Fischer scientific) were coated with anti-human IgA_2_ antibody (Miltenyi) 1μg/ml in Bicarbonate buffer, and incubated for 1.5h at room temperature. After washing with PBS-Tween (PBST) (Sigma), blocking was done using PBST with 3% BSA during 1.5h at room temperature. Before being added to the plate, plasma and serum samples were diluted 1:100 in PBST-BSA, while tissue supernatant was diluted 1:3 in PBST-BSA. Serial dilutions (1:3) were made through the plate, and samples were incubated overnight at 4°C. Plates were washed 3x with PBST, and secondary anti-IgA_2_-HRP (Southern Biotech) antibody was added and incubated 1.5h at room temperature. Finally, plates were washed with PBST and TMB substrate was added. Absorbance was measured at 450nm using a plate reader.

### Tissue culture

During colonoscopy, biopsies were taken with forceps directly into ice-cold RPMI with 10x Antibiotic-Antimycotic (RPMI-AA) (Gibco), and transported to the lab on ice. Under sterile conditions, biopsies were rinsed on a cell strainer using RPMI-AA 10x. Biopsies were placed in pairs into 48-well plates with 1 ml RPMI-AA 10x per well, and kept from this point at 37°C with 5% CO_2_. Culture medium was replaced every 2 days using pre-warmed RPMI-AA 2x. Recovered culture medium was centrifuged 10min at 14000G and stored at - 80°C.

### Bacterial flow cytometry

Fecal pellets were dissolved in PBS at 100 mg/ml by vortexing. Fecal slurry was centrifuged 10min at 50G to remove large particles. Supernatant was filtered through a 40nm cell strainer and cells were then washed by centrifugation at 9000G and resuspension in washing buffer (PBS, 1% BSA, 2mM EDTA). Blocking was done using PBS +1% BSA +20% mouse serum, for 20min at 4°C. Cells were stained with anti-human IgA2-PE and anti-human IgA-APC for 30min at 4°C, before washing three times with washing buffer. Samples were stained with SYBR green before analysis by flow cytometry.

### Immune globulin measurements

For patient CD068, serum immune globulins were IgG=1,435 (nl 603 - 1,613 mg/dl); IgA=387 (nl 90-386 mg/dl); and IgM= 197 (l n20 - 172 mg/dl). IgE=13 (6-495IU/ml). In terms of IgG subclasses, IgG1- 753 (248 - 810 mg/dL) IgG2 = 464(130 - 555 mg/dL) IgG3 = 108 (15 - 102 mg/dL); IgG4 = 52 (2 - 96 mg/dl). His IgA1 = 391.6 (73.2 - 301.2 mg/dL) mg /dl, and IgA2 = <0.5 (13.4 - 97.9 mg/dL).

### Genetic Primary Immunodeficiency Panel

A primary immune deficiency panel of 407 genes was obtained with patient’s consent (Invitae Corp, San Francisco CA).

### In vitro B cell stimulation

For B cell activation, PBMCs were cultured in RPMI media (Gibco) supplemented with 10% FBS, the presence of CD40L (1μg/ml) (InvivoGen) plus IL-21 (100 ng/ml) (BioLegend) for 10 days. Cells were kept in 24-well plates at 37°C with 5% CO_2_. After 10 days of culture, cells were analyzed by Flow cytometry, as indicated before.

---

**Figure S1.**
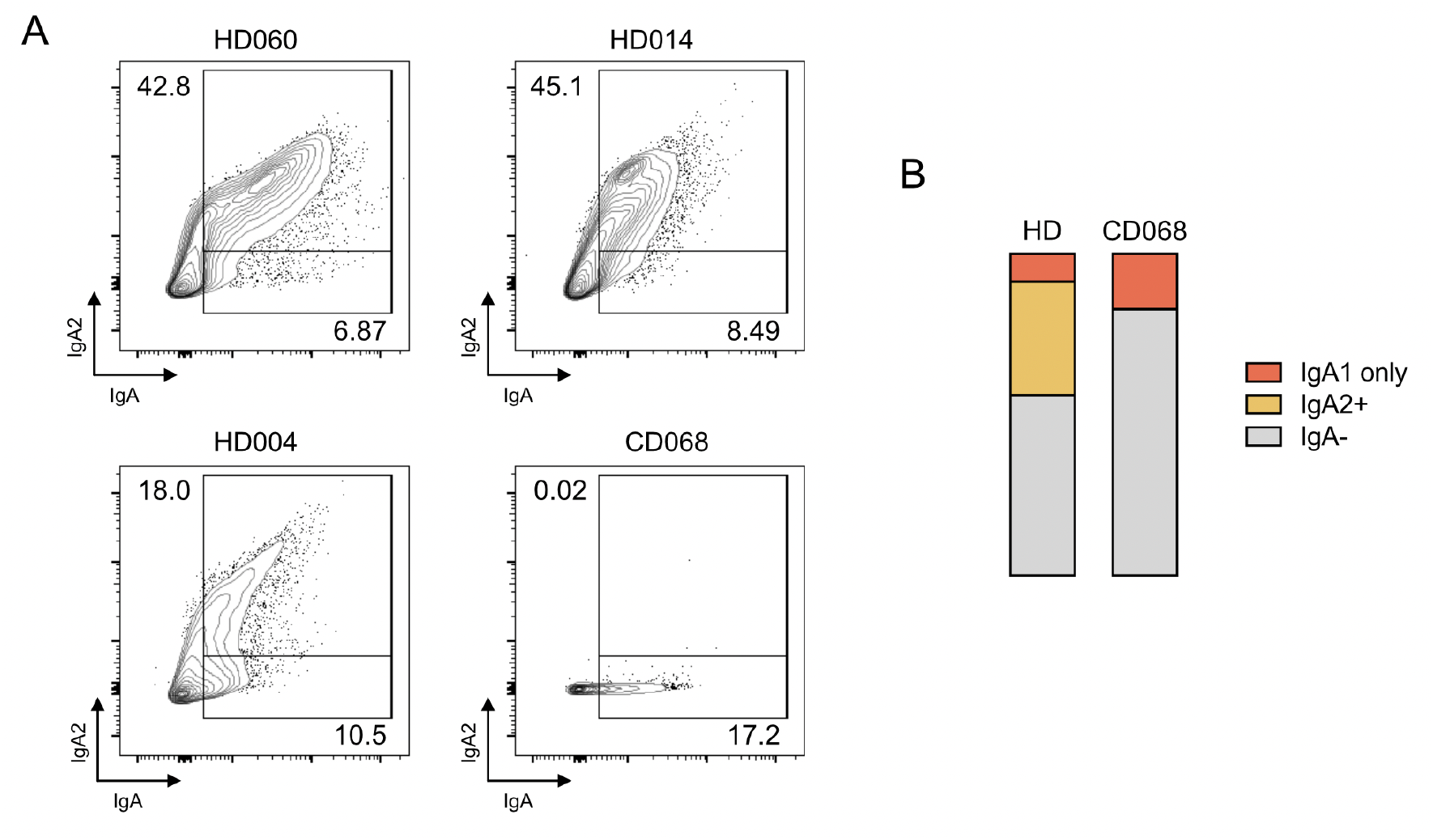
Profile of IgA coating on fecal microbiota. (A) Flow cytometric analysis of IgA+ and IgA_2_+ microbiota from healthy donors and patient CD068. Cells were previously gated for SYBR green positivity. Frequency of IgA_2_+ and IgA_1_ only are indicated. (B) Average proportions of IgA_1_ only, IgA_2_+ and IgA-cells are indicated for HDs and patient CD068.

**Figure S2.**
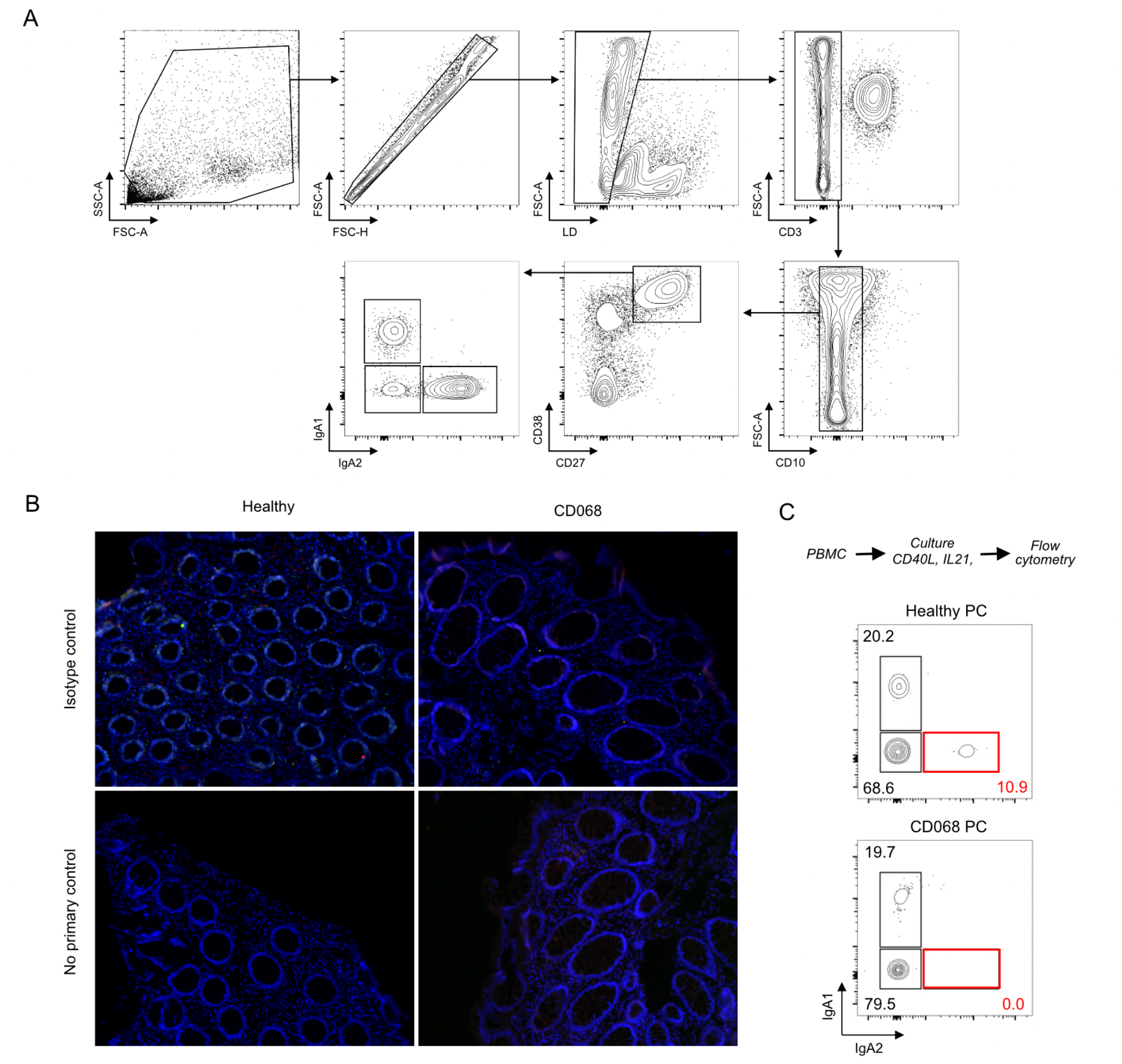
Unknown B cell-intrinsic defect in a patient with selective IgA_2_ deficiency associated with small intestinal CD. (A) Gating strategy used to identify viable CD3^-^CD10^−^CD27^+^CD38^high^ PCs within colonic lamina propria mononuclear cells. (B) Immunofluorescence analysis of isotype controls or non-primary IgA_1_ and IgA_2_ controls in colonic tissue sections from a representative HD (left) or patient CD068. (C) Flow cytometric staining of IgA_1_ and IgA_2_ on PCs from PBMCs stimulated *in vitro* for 10 days with CD40L and IL-21. PCs were gated as in (A).

**Table S1.**
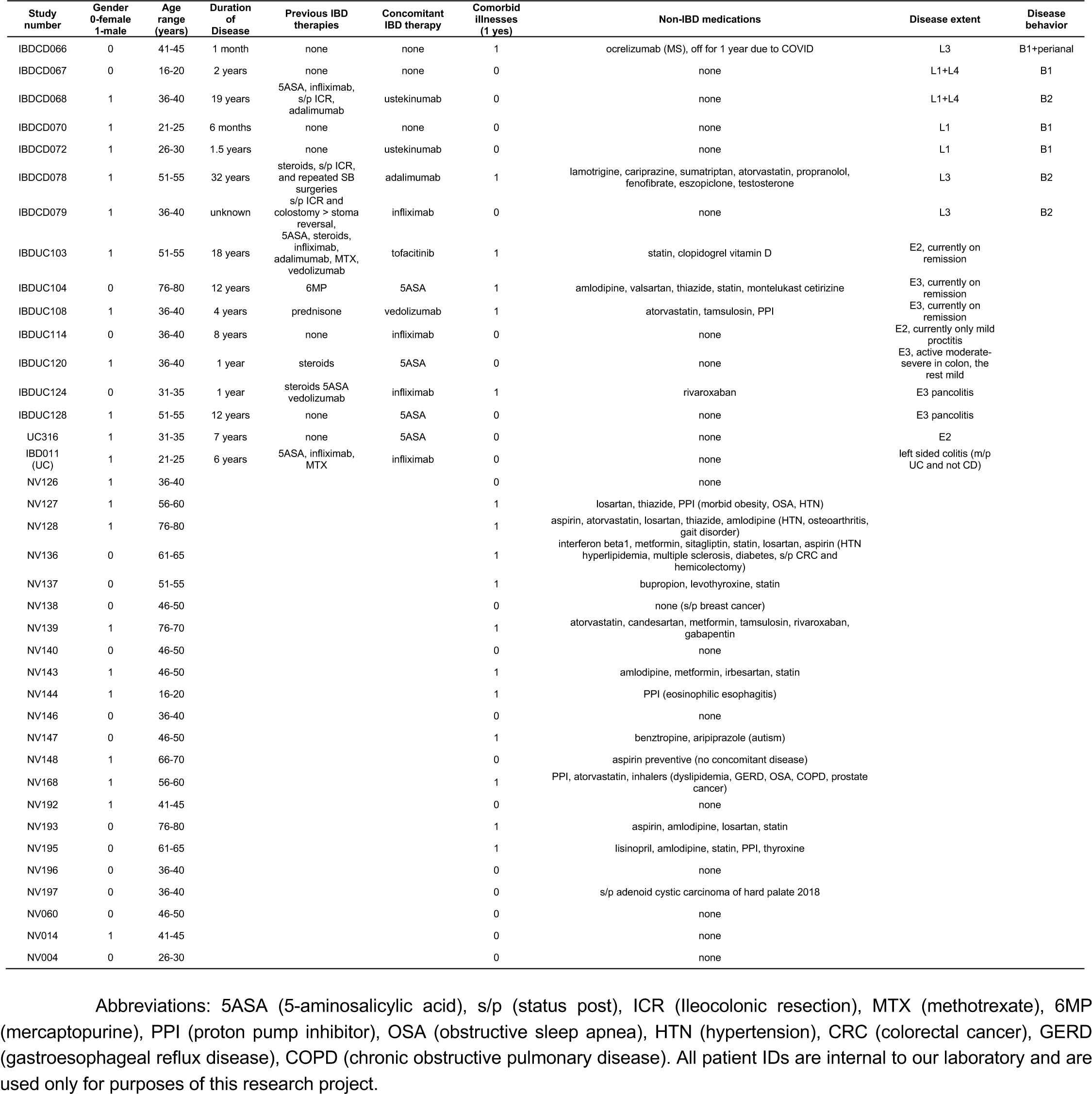
Patient characteristics (where flow cytometry was performed)

**Table S2.**
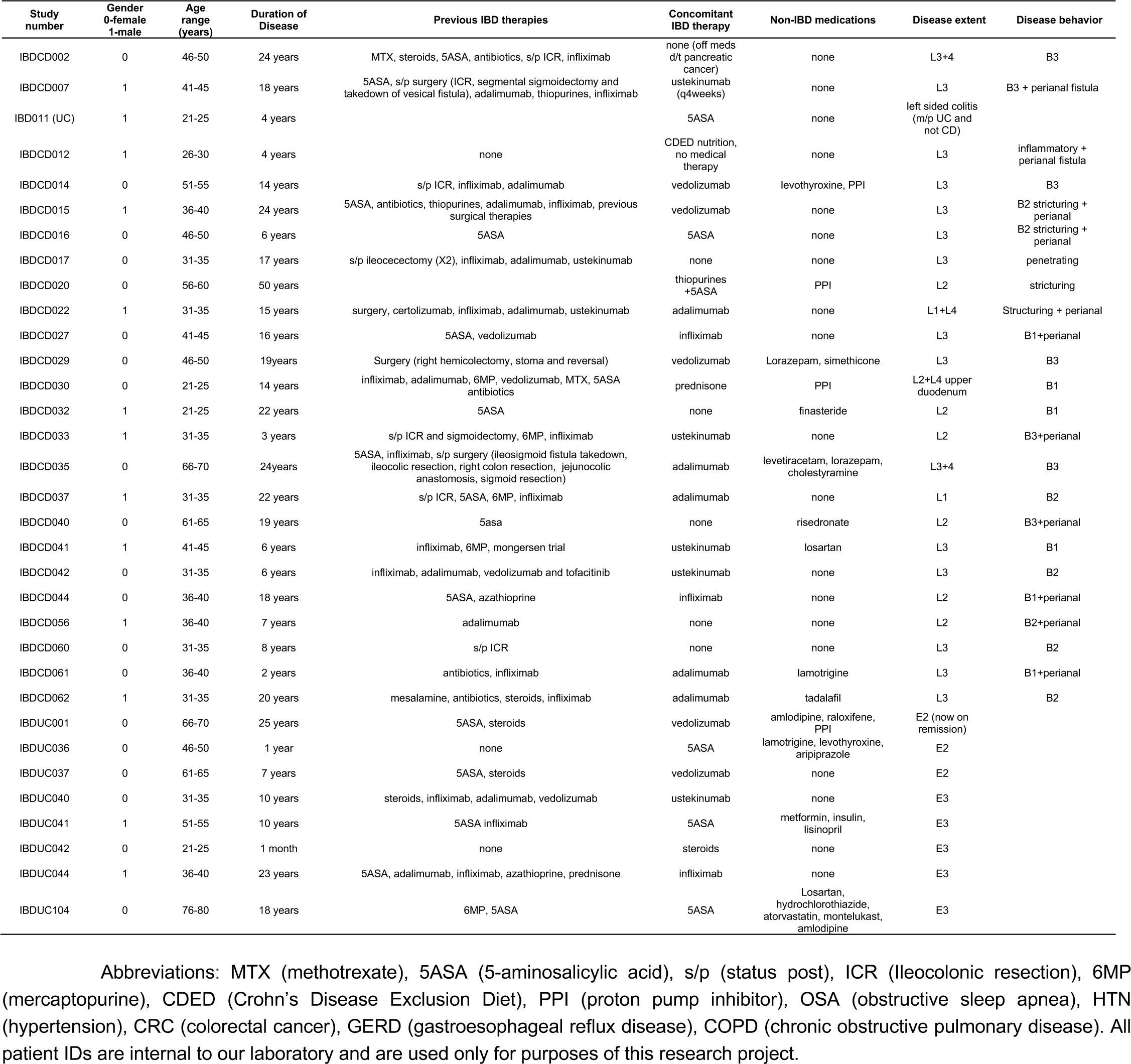
Patient characteristics (where plasma was used for immunoglobin quantification)

**Table S3.** Antibodies used in the study.

| Flow cytometry |  |  |  |  |
| --- | --- | --- | --- | --- |
| Marker | Fluorochrome | Supplier | Catalog # | Clone |
| CD19 | PE Cy7 | Biologend | 363012 | SJ25C1 |
| CD27 | PerCP Cy5.5 | Biologend | 302819 | O323 |
| CD10 | APC Cy7 | Biologend | 312212 | HI10a |
| CD38 | APC | Biologend | 356606 | HB-7 |
| IgD | PB | Biologend | 348223 | IA6-2 |
| IgM | AF700 | Biologend | 314538 | MHM-88 |
| CD45 | PE Dazzle 594 | Biologend | 304052 | HI30 |
| CD3 | BV605 | Biologend | 300460 | UCHT1 |
| IgA1 | A488 | Southern Biotech | 9130-30 | B3506B4 |
| IgA2 | PE | Miltenyi | 130-100-316 | IS11-21E11 |
| Immunofluorescence |  |  |  |  |
| Marker | Fluorochrome | Supplier | Catalog # | Clone |
| rabbit anti-human IgA1 | pure | Abcam | ab193187 | RM124 |
| mouse anti-human IgA2 | pure | Miltenyi | 130-093-081 | IS11-21E11 |
| goat anti-human IgA | pure | Abcam | ab98550 | polyclonal |
| goat anti-mouse | AF594 | Abcam | ab150116 | polyclonal |
| goat anti-rabbit | AF488 | Abcam | ab150077 | polyclonal |
| donkey anti-goat | AF594 | Abcam | ab150132 | polyclonal |
| donkey anti-rabbit | AF488 | Abcam | ab150073 | polyclonal |

## Notes

### Competing Interest Statement

The authors have declared no competing interest.

### Author Declarations

This study was approved by the institutional review board of the Icahn School of Medicine at Mount Sinai (17-01304).

## References

1. Chen K, Magri G, Grasset EK, Cerutti A. Rethinking mucosal antibody responses: IgM, IgG and IgD join IgA. Nat Rev Immunol. 2020 Jul;20(7):427–441.

2. van Loghem E, Zegers BJ, Bast EJ, Kater L. Selective deficiency of immunoglobulin A2. J Clin Invest. 1983 Dec;72(6):1918–23.

3. Ozawa N, Shimizu M, Imai M, Miyakawa Y, Mayumi M. Selective absence of immunoglobulin A1 or A2 among blood donors and hospital patients. Transfusion. 1986 Jan-Feb;26(1):73–6.

4. Engström PE, Norhagen G, Bottaro A, Carbonara AO, Lefranc G, Steinitz M, Söder PO, Smith CI, Hammarström L. Subclass distribution of antigen-specific IgA antibodies in normal donors and individuals with homozygous C alpha 1 or C alpha 2 gene deletions. J Immunol. 1990 Jul 1;145(1):109–16.

5. Mesin L, Di Niro R, Thompson KM, Lundin KE, Sollid LM. Long-lived plasma cells from human small intestine biopsies secrete immunoglobulins for many weeks in vitro. J Immunol. 2011 Sep 15;187(6):2867–74.

6. Ludvigsson JF, Neovius M, Hammarström L. Association between IgA deficiency & other autoimmune conditions: a population-based matched cohort study. J Clin Immunol. 2014 May;34(4):444–51.

7. Chen K, Magri G, Grasset EK, Cerutti A. Rethinking mucosal antibody responses: IgM, IgG and IgD join IgA. Nat Rev Immunol. 2020 Jul;20(7):427–441.

8. van Loghem E, Zegers BJ, Bast EJ, Kater L. Selective deficiency of immunoglobulin A2. J Clin Invest. 1983 Dec;72(6):1918–23.

9. Ozawa N, Shimizu M, Imai M, Miyakawa Y, Mayumi M. Selective absence of immunoglobulin A1 or A2 among blood donors and hospital patients. Transfusion. 1986 Jan-Feb;26(1):73–6.

10. Engström PE, Norhagen G, Bottaro A, Carbonara AO, Lefranc G, Steinitz M, Söder PO, Smith CI, Hammarström L. Subclass distribution of antigen-specific IgA antibodies in normal donors and individuals with homozygous C alpha 1 or C alpha 2 gene deletions. J Immunol. 1990 Jul 1;145(1):109–16.

11. Mesin L, Di Niro R, Thompson KM, Lundin KE, Sollid LM. Long-lived plasma cells from human small intestine biopsies secrete immunoglobulins for many weeks in vitro. J Immunol. 2011 Sep 15;187(6):2867–74.

12. Ludvigsson JF, Neovius M, Hammarström L. Association between IgA deficiency & other autoimmune conditions: a population-based matched cohort study. J Clin Immunol. 2014 May;34(4):444–51.

13. Uzzan M, Martin JC, Mesin L, Livanos AE, Castro-Dopico T, Huang R, Petralia F, Magri G, Kumar S, Zhao Q, Rosenstein AK, Tokuyama M, Sharma K, Ungaro R, Kosoy R, Jha D, Fischer J, Singh H, Keir ME, Ramamoorthi N, Gorman WEO, Cohen BL, Rahman A, Cossarini F, Seki A, Leyre L, Vaquero ST, Gurunathan S, Grasset EK, Losic B, Dubinsky M, Greenstein AJ, Gottlieb Z, Legnani P, George J, Irizar H, Stojmirovic A, Brodmerkel C, Kasarkis A, Sands BE, Furtado G, Lira SA, Tuong ZK, Ko HM, Cerutti A, Elson CO, Clatworthy MR, Merad M, Suárez-Fariñas M, Argmann C, Hackney JA, Victora GD, Randolph GJ, Kenigsberg E, Colombel JF, Mehandru S. Ulcerative colitis is characterized by a plasmablast-skewed humoral response associated with disease activity. Nat Med. 2022 Apr;28(4):766–779.

14. Castro-Dopico T, Dennison TW, Ferdinand JR, Mathews RJ, Fleming A, Clift D, Stewart BJ, Jing C, Strongili K, Labzin LI, Monk EJM, Saeb-Parsy K, Bryant CE, Clare S, Parkes M, Clatworthy MR. Anti-commensal IgG Drives Intestinal Inflammation and Type 17 Immunity in Ulcerative Colitis. Immunity. 2019 Apr 16;50(4):1099-1114.e10.

15. Kett K, Brandtzaeg P. Local IgA subclass alterations in ulcerative colitis and Crohn’s disease of the colon. Gut. 1987 Aug;28(8):1013–21.

16. Kett K, Brandtzaeg P, Fausa O. J-chain expression is more prominent in immunoglobulin A2 than in immunoglobulin A1 colonic immunocytes and is decreased in both subclasses associated with inflammatory bowel disease. Gastroenterology. 1988 Jun;94(6):1419–25.

17. Boland BS, He Z, Tsai MS, Olvera JG, Omilusik KD, Duong HG, Kim ES, Limary AE, Jin W, Milner JJ, Yu B, Patel SA, Louis TL, Tysl T, Kurd NS, Bortnick A, Quezada LK, Kanbar JN, Miralles A, Huylebroeck D, Valasek MA, Dulai PS, Singh S, Lu LF, Bui JD, Murre C, Sandborn WJ, Goldrath AW, Yeo GW, Chang JT. Heterogeneity and clonal relationships of adaptive immune cells in ulcerative colitis revealed by single-cell analyses. Sci Immunol. 2020 Aug 21;5(50): eabb4432.

18. Bunker JJ, Flynn TM, Koval JC, Shaw DG, Meisel M, McDonald BD, Ishizuka IE, Dent AL, Wilson PC, Jabri B, Antonopoulos DA, Bendelac A. Innate and Adaptive Humoral Responses Coat Distinct Commensal Bacteria with Immunoglobulin A. Immunity. 2015 Sep 15;43(3):541–53.

19. Sterlin D, Fadlallah J, Adams O, Fieschi C, Parizot C, Dorgham K, Rajkumar A, Autaa G, El-Kafsi H, Charuel JL, Juste C, Jönsson F, Candela T, Wardemann H, Aubry A, Capito C, Brisson H, Tresallet C, Cummings RD, Larsen M, Yssel H, von Gunten S, Gorochov G. Human IgA binds a diverse array of commensal bacteria. J Exp Med. 2020 Mar 2;217(3):e20181635.

20. Nagaishi T, Watabe T, Kotake K, Kumazawa T, Aida T, Tanaka K, Ono R, Ishino F, Usami T, Miura T, Hirakata S, Kawasaki H, Tsugawa N, Yamada D, Hirayama K, Yoshikawa S, Karasuyama H, Okamoto R, Watanabe M, Blumberg RS, Adachi T. Immunoglobulin A-specific deficiency induces spontaneous inflammation specifically in the ileum. Gut. 2022 Mar;71(3):487–496.

21. Zegers BJ, van Loghem E, Out T, et al. Familial deficiency of immunoglobulin IgA2. Monogr Allergy. 1986;20:195–202.

22. Oliviero S, DeMarchi M, Bast BJ, Zegers BJ, van Loghem E, de Lange G, Carbonara O. Molecular analysis of a case of IgA2 deficiency. J Immunogenet. 1986 Feb;13(1):3–9. doi: 10.1111/j.1744-313x.1986.tb01077.x.

23. Bottaro A, De Marchi M, De Lange G, Boccazzi C, Caldesi F, Gallina R, Carbonara AO. New types of multiple and single gene deletions in the human IgCH locus. Immunogenetics. 1989;29(1):44–8. doi: 10.1007/BF02341612. Erratum in: Immunogenetics 1989;29(3):224

